# Novel biomarkers of habitual alcohol intake and associations with risk of pancreatic and liver cancers and liver disease mortality

**DOI:** 10.1101/2021.03.12.20224451

**Authors:** Erikka Loftfield, Magdalena Stepien, Vivian Viallon, Laura Trijsburg, Joseph Rothwell, Nivonirina Robinot, Carine Biessy, Ingvar A. Bergdahl, Stina Bodén, Matthias B. Schulze, Manuela Bergman, Elisabete Weiderpass, Julie A. Schmidt, Raul Zamora-Ros, Therese H. Nøst, Torkjel M Sandanger, Emily Sonestedt, Bodil Ohlsson, Verena Katzke, Rudolf Kaaks, Fulvio Ricceri, Anne Tjønneland, Christina C. Dahm, Maria-Jose Sánchez, Antonia Trichopoulou, Rosario Tumino, María-Dolores Chirlaque, Giovanna Masala, Eva Ardanaz, Roel Vermeulen, Paul Brennan, Demetrius Albanes, Stephanie J. Weinstein, Augustin Scalbert, Neal D. Freedman, Marc J. Gunter, Mazda Jenab, Rashmi Sinha, Pekka Keski-Rahkonen, Pietro Ferrari

## Abstract

**Background:** Alcohol is an established risk factor for several cancers, but modest alcohol-cancer associations may be missed due to measurement error in self-reported assessments. The identification of biomarkers of habitual alcohol intake may enhance evidence on the role of alcohol in cancer onset.

**Methods:** Untargeted metabolomics was used to identify metabolites correlated with habitual alcohol intake in a discovery dataset from the European Prospective Investigation into Cancer and Nutrition (EPIC; n=454). Significant correlations were replicated in independent datasets of controls from case-control studies nested within EPIC (n=281) and the Alpha-Tocopherol, Beta-Carotene Cancer Prevention (ATBC; n=438) study. Conditional logistic regression was used to estimate odds ratios (OR) and 95% confidence intervals for associations of alcohol-associated metabolites and self-reported alcohol intake with risk of pancreatic cancer, hepatocellular carcinoma (HCC), liver cancer, and liver disease mortality in the contributing studies.

**Results:** Two metabolites displayed a dose-response association with alcohol intake: 2-hydroxy-3-methylbutyric acid and an unidentified compound (*m/z(+)*:231.0839). A 1-SD increase in log_2_-transformed levels of 2-hydroxy-3-methylbutyric acid was associated with risk of HCC (OR=2.14; 1.39-3.31) and pancreatic cancer (OR=1.65; 1.17-2.32) in EPIC and liver cancer (OR=2.00; 1.44-2.77) and liver disease mortality (OR=2.16; 1.63-2.86) in ATBC. Conversely, a 1-SD increase in log_2_-transformed questionnaire-derived alcohol intake was not associated with HCC or pancreatic cancer in EPIC or liver cancer in ATBC but was associated with liver disease mortality (OR=2.19; 1.60-2.98) in ATBC.

**Conclusions:** 2-Hydroxy-3-methylbutyric acid is a candidate biomarker of habitual alcohol intake that may advance the study of alcohol and cancer risk in population-based studies.

---

In 2016, an estimated 2.8 million deaths, corresponding to 6.8% and 2.2% of age-standardized deaths in men and women, respectively, were attributed to alcohol use worldwide [1]. Excessive alcohol consumption is an established risk factor for many acute and chronic health conditions [2], including cancers of the upper aerodigestive tract, female breast, liver, colon and rectum [3]. However, the relationship of alcohol, particularly light-to-moderate alcohol consumption, with other cancer sites remains controversial [4].

Self-reported alcohol intake is, like other dietary factors, prone to underreporting [5]. Although the extent and distribution of exposure misclassification is unknown [6], it is likely that observed associations between alcohol use and disease risk in prospective studies are attenuated and that estimates of alcohol-attributable death and disease are underestimated. Biomarkers of liver function and oxidative stress are used to study alcohol-related liver injury and alcoholic liver disease (ALD) [7, 8], but most alcohol consumers, particularly light-to-moderate consumers, will never manifest ALD. There are also biomarkers of recent (e.g., ethyl glucuronide) and heavy alcohol use (e.g., carbohydrate deficient transferrin and phosphatidylethanol (PEth)) [9-11]. However, biomarkers of habitual alcohol use, including light-to-moderate drinking, are needed to better assess alcohol exposure in epidemiological studies and to improve risk estimates for diseases including cancer where modest associations may exist.

Metabolomics is a powerful tool for discovering dietary biomarkers. When used in an untargeted mode, it can detect a wide range of compounds in biological samples including metabolites formed during digestion, metabolism and microbial fermentation [12, 13], making it well-suited for discovering novel biomarkers of exposure or response to habitual alcohol consumption. Herein we applied a multistep design, using untargeted metabolomics and independent discovery and replication datasets, to identify serum metabolites associated with habitual alcohol consumption among free-living individuals with a wide range of intake. We then estimated the associations of these candidate alcohol biomarkers with risk of pancreatic cancer, liver cancers, and liver disease mortality in the European Prospective Investigation into Cancer and Nutrition (EPIC) study and the Alpha-Tocopherol, Beta-Carotene Cancer Prevention Study (ATBC).

## METHODS

### Study design

#### EPIC Study

EPIC recruitment and study procedures, including dietary assessment methods and blood collection are described extensively elsewhere [14]. Briefly, EPIC is a large cohort study of over half a million men and women recruited between 1992 and 2000 in 23 European centers. Diet, including average daily alcohol intake, over the 12 months before enrolment was assessed by validated country-specific food frequency questionnaires (FFQ) designed to capture local dietary habits with high compliance. Country-specific alcohol intake was calculated based on the estimated average glass volume and ethanol content for wine, beer, cider, sweet liquor, distilled spirits, or fortified wines, using information collected in standardized 24-hr dietary recalls from a subset of the cohort [15]. The correlation between alcohol intake estimated by FFQ and 24-hour dietary recall was 0.79 [16]. Blood samples were collected and stored at -196°C under liquid nitrogen at the International Agency for Research on Cancer (IARC) for all countries except Sweden (−80°C freezers), and Denmark (−150°C, nitrogen vapor).

Our study included a discovery and two replication datasets (**Figure 1**). The discovery set (n=454) was nested in the EPIC cross-sectional study [17, 18]. The first replication set included control subjects from two EPIC nested case-control studies of hepatocellular carcinoma (HCC; n=129) and pancreatic cancer (n=152) with untargeted metabolomics data [19-21]. Non-metastatic incident HCC (n=129) and pancreatic cancer (n=152) cases, were matched 1:1 with cancer-free controls on study center, sex, age at blood collection (□±□1 year), date (□±□6 months) and time of the day (□±□2□h) of blood collection, fasting status, and, for women, exogenous hormone use. Follow-up was based on a combination of methods, including health insurance records, registries and active follow-up [14]. Approval for the EPIC study was obtained from the IARC ethics review board (Lyon, France) and local review bodies of participating institutions.

**Figure 1.**
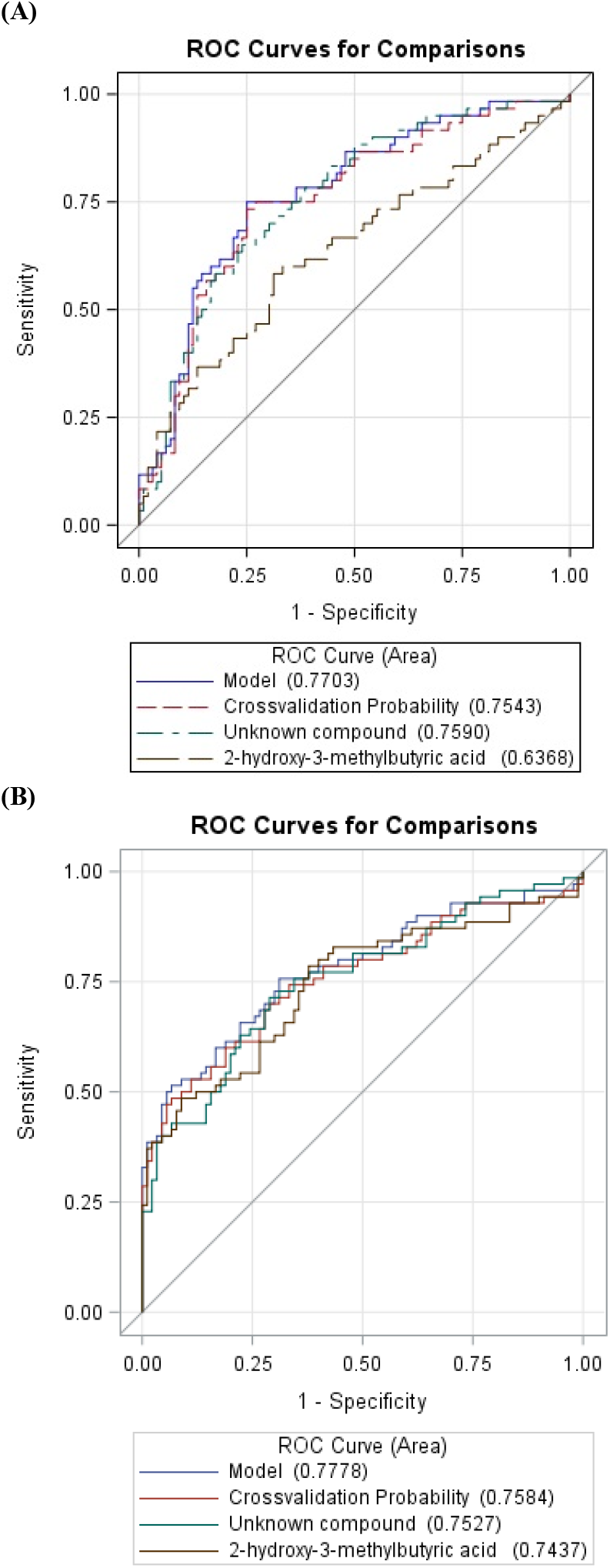
Flowchart of the study, displaying features and samples size of the discovery and Replication I (EPIC) and II (ATBC) sets (blue box), as well as of the aetiological components in nested-case-control studies (red box).

#### ATBC Study

The second replication set included two nested case-control studies in the ATBC cohort of male Finnish smokers [22]. In ATBC, participants reported on demographics, lifestyle, and medical history via questionnaires and donated a fasting serum sample at baseline, which was stored at -70°C. Participants were passively followed during the post-intervention period via linkage with the Finnish Cancer Registry and death registry. Liver cancer (n=229) and liver disease mortality (n=248) cases were individually matched 1:1 with controls, selected by incidence density sampling, on baseline age (+/-5 years) and serum draw date (+/-30 days) [23]. For this study, we excluded cases and controls with missing data on alcohol intake (n=72) and those with samples that failed laboratory analysis (n=14) resulting in an analytic sample of n=438 controls for cross-sectional analysis and 192 and 199 complete case-control sets for prospective analyses of liver cancer incidence and liver disease mortality, respectively. Approval for the ATBC study was obtained from the Institutional Review Boards of National Cancer Institute (Bethesda, Maryland), and the National Public Health Institute of Finland. EPIC and ATBC studies were conducted according to the guidelines of the Declaration of Helsinki; all participants provided written informed consent.

### Metabolomics analyses

Sample analysis, data pre-processing, matching of features across datasets, and compound identification are desribed in detail in the

### Supplementary Methods

Briefly, all samples were analyzed by the same laboratory at IARC with a UHPLC-QTOF-MS system (1290 Binary LC system, 6550 QTOF mass spectrometer; Agilent Technologies, Santa Clara, CA) using reversed phase chromatography and electrospray ionization. Raw data were processed using Agilent MassHunter Qualitative analysis B.06.00, ProFinder B.08.00, and Mass Profiler Professional B.12.1 software with Agilent’s recursive feature finding procedure. The *m/z* values of the features of interest were searched against the Human Metabolome Database (HMDB) [24] and METLIN [25].Compound identity was confirmed by comparison of chemical standards and representative samples.

### Statistical analyses

We used an integrated workflow for metabolomics data analysis [26]. Features detected in <50% of the discovery set samples abd ackground features, (i.e., feature intensities present in all blanks with ratio of geometric mean intensities of non-blank:blank samples <5) were excluded. Feature intensities were log_2_-transformed. Study participants with >50% missing features and those identified as outliers by a PCA-based approach were excluded [27]. Missing values were imputed within each plate by a K-nearest neighbours method, with K=10 [28]. Last, feature intensities measured across plates within any single batch were normalised by applying a random forest-based approach to correct for unwanted variation [29]. In the EPIC discovery set and replication sets, these steps were applied on feature matrices acquired in positive and negative modes separately. In ATBC, these steps were applied on each batch.

In the discovery and replication sets, alcohol intake (g/day) was adjusted for age, sex, country (in EPIC only), body mass index (BMI, kg/m^2^), smoking status and intensity, coffee consumption (g/day, log-transformed) via the residual method in linear regression models. Coffee drinking and coffee-associated metabolites have been strongly associated with risk of liver cancer and liver disease mortality in ATBC [30, 31]; for consistency, coffee drinking was considered a potential confounder across discovery and replication sets. Residuals for feature intensities were also adjusted for well plate number within the analytical batch, position within the plate (row and column indexes), and the study (EPIC replication) or batch indicator (ATBC replication) as random effects. We used the principal component partial-R^2^ (PC-PR2) method [32] to quantify the contribution of alcohol and potential confounders to the variability of the 67 features intensities that were statistically significantly associated with alcohol intake in the discovery set [33].

We calculated Pearson correlation coefficients using the residuals for alcohol intake and for feature intensities; correlations with a false discovery rate (FDR)-corrected p-value<0.05 were considered statistically significant, and these *f*_1_ features were carried forward for replication. In the first (EPIC) replication step, *f* _1_ residual-adjusted correlation coefficients were computed and corrected by the more conservative Bonferroni method. The *f* _2_ correlations with a p-value <0.05/*f* _1_ were considered statistically significant and carried forward to the second replication step in ATBC, again using the residuals for alcohol intake and for feature intensities. The linearity of the association between standardized residuals of 2-hydroxy-3-methylbutyric acid and of alcohol intake was evaluated with cubic regression splines with 5 knots [34], by comparing the log-likelihood of models with and without the non-linear terms to a chi-distribution with 2 degrees of freedom.

The EPIC replication set was used to define high- (quartile 4: alcohol intake >33.1 and >12.3 g/day in men and women, respectively)) and low-consumers (quartile 1: alcohol intake >0.1 but <4.9 and <1.1 g/day in men and women, respectively). We used logistic regression to estimate the area under receiver operating characteristics (AUROC) curves [35] and evaluated the predictive accuracy of the residuals of each main feature (i.e., candidate biomarkers) to discriminate high-consumers from low-consumers for metabolites that replicated across studies. We used the leave-one-out cross validation scheme, to mitigate issues related to over fitting [36].

We estimated odds ratios (OR) and 95% confidence intervals (95% CI) for candidate features and HCC and pancreatic cancer in EPIC and liver cancer and fatal liver disease in ATBC using conditional logistic regression models. In crude (conditioned on the matching criteria only) and multivariable models, adjusting for potential confounders, feature intensities were log_2_-transformed, centered and scaled (i.e., mean=0, standard deviation=1) to ensure comparability of OR across different endpoints.

All statistical analyses were performed using the Statistical Analysis Software, release 9.4 (SAS Institute Inc., Cary, NC, USA) and R version 3.6.0 [37].

## RESULTS

### Population characteristics

Baseline participant characteristics are presented in **Table 1**. In the EPIC discovery set, most participants were women (57.5%) and never (52.2%) or former (26.4%) smokers. In the EPIC replication set, there was a higher percentage of men (52.7%) and a lower percentage of never smokers (46.2%). In the ATBC replication set, all participants were Finnish men and current smokers. Median alcohol intake was 10.0 g/day, 6.6 g/day, and 11.5 g/day in the EPIC discovery, EPIC and ATBC replication sets, respectively.

**Table 1.** Descriptive statistics of the discovery and replication sets.

|  | **EPIC Discovery<sup>1</sup>** | **EPIC Replication<sup>2</sup>** | **ATBC Replication<sup>3</sup>** |
| --- | --- | --- | --- |
|  | **(n=454)** | **(n=281)** | **(n=438)** |
| Men (%) | 42.5 | 52.7 | 100 |
| BMI (median kg/m<sup>2</sup>; 10-90<sup>th</sup> %) | 25.8 (20.9-31.6) | 26.6 (20.7-34.1) | 26.2 (22.5-31.3) |
| Age (median years; 10-90<sup>th</sup> %) | 55.2 (42.5-63.9) | 59.4 (49.0-68.6) | 56.0 (51.0-63.0) |
| Smoking status (%) |  |  |  |
| Current | 18.5 | 19.2 | 100 |
| Former | 26.4 | 33.5 |  |
| Never | 52.2 | 46.2 |  |
| Unknown | 2.9 | 1.1 |  |
| Smoking intensity (median cig/day; 10-90<sup>th</sup> %) | 11.5 (2-26) | 15 (4-30) | 20 (10-30) |
| Country (%) |  |  |  |
| France | 14.5 | 0.4 |  |
| Italy | 34.8 | 18.5 |  |
| Spain | - | 10.0 |  |
| United Kingdom | - | 17.1 |  |
| The Netherlands | - | 10.3 |  |
| Greece | 12.3 | 10.7 |  |
| Germany | 38.3 | 24.9 |  |
| Denmark | - | 8.2 |  |
| Finland | - | - | 100 |
| Alcohol non-drinkers (%)<sup>4</sup> | 8 | 14 | 9 |
| Alcohol intake (median g/day; 10<sup>th</sup>-90<sup>th</sup> %) |  |  |  |
| Men | 21.4 (1.3-50.4) | 14.9 (1.0-51.7) | 11.5 (0.2-42.1) |
| Women | 5.2 (0.02-24.9) | 2.0 (0.01-23.3) | -- |
| Coffee intake (median g/day; 10<sup>th</sup>-90<sup>th</sup> %) | 146.3 (21.4, 580.2) | 190 (3, 857) | 550 (220-1,100) |

<sup>1</sup>EPIC cross-sectional sample;<sup>2</sup>Controls from both liver and pancreatic cancer EPIC nested case-control studies;<sup>3</sup>Controls from liver cancer and liver disease mortality ATBC nested case-control studies excluding those with missing data on alcohol intake;<sup>4</sup>Alcohol non-drinkers are considered as those with alcohol intake $\leq 0.1$ g/day.

### Biomarker discovery analysis

After excluding participant samples identified as outliers or as having too many missing values, the final discovery set comprised 451 and 452 study participants in positive and negative ionization mode datasets, respectively. The final EPIC replication set comprised 271 and 277 study participants in positive and negative ionization datasets, respectively. Residuals of 205 features in the discovery set were significantly correlated with residuals of alcohol intake (163 features in positive and 42 features in negative ionization mode; **Figure 1)**, with correlation coefficients ranging from - 0.29 to 0.50 in log-log plots (**Table S1**).

Of the 205 features in the discovery set, 51 features in positive and 16 features in negative ionization mode (*f* _1_=67) matched by mass and retention time with equivalent features in the EPIC replication set, and PC-PR2 analyses showed that alcohol intake explained >7% of variability in the feature intensities (*f* _1_=67; **Figure 2**). Residuals of *f*_2_=10 features were statistically significantly correlated with residuals of alcohol intake (**Table 2**). The first two features corresponded to a compound that could not be unequivocally identified, but had an identical mass, isotope pattern, ion formation (mostly [M+Na]^+^ and [M+HCOOH-H]^-^) and retention time to ethyl glucoside (HMDB0029968) [38]. However, chromatograms (**Supplementary Methods**) indicated a lack of specificity, and although fragmentation of the [M+Na]^+^ ion could not be induced, our results suggest the unknown is a combination of ethyl-α-D-glucoside, ethyl-β-D-glucoside, and an additional structural isomer. The remaining eight features corresponded to a single compound, which was confirmed by comparison with an authentic standard as 2-hydroxy-3-methylbutyric acid (HMDB0000407).

**Figure 2.**
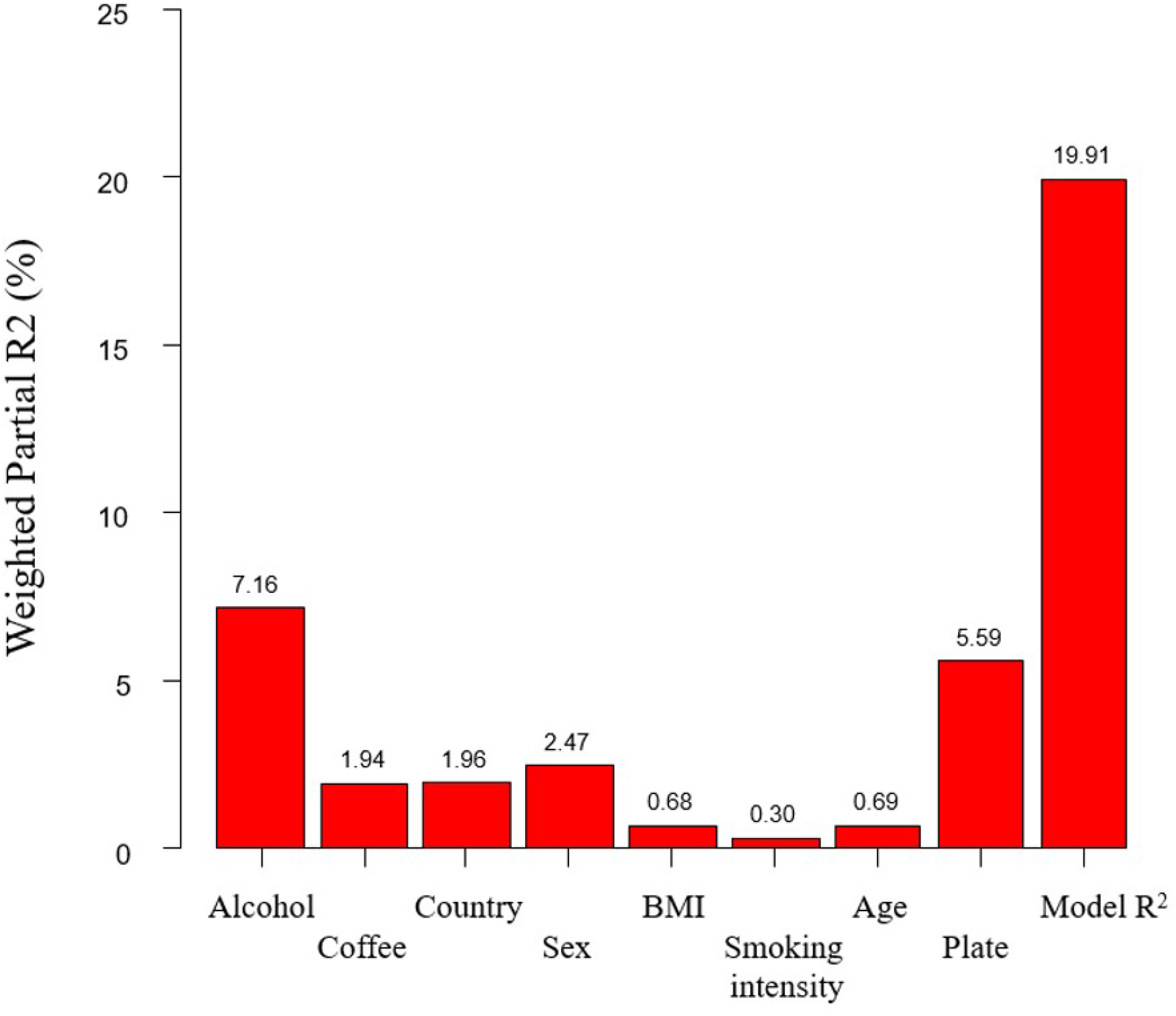
PC-PR2 (Principal Component Partial R^2^) analysis to quantify the contribution of potential confounder variables to the variability of the set of *f*_1_=67 feature intensities that were statistically significantly associated to alcohol intake in the discovery set.

**Table 2.** Feature-specific intensity and reproducibility (coefficient of variation=CV) in quality control (QC) samples, and adjusted Pearson correlation coefficients (r) with between alcohol intake in the discovery and replication sets.

| *m/z*<sup>4</sup> | RT<sup>5</sup><br>(min) | Method | Associated metabolite | QC samples<sup>1</sup><br>(n=38) |  | EPIC Discovery<br>(n=454)<sup>2</sup> |  |  | EPIC Replication<br>(n=281)<sup>3</sup> |  | ATBC Replication<br>(n=438) |  |
| --- | --- | --- | --- | --- | --- | --- | --- | --- | --- | --- | --- | --- |
|  |  |  |  | Mean<br>intensity | CV<br>(%) | r | p-value | q-value<sup>6</sup> | r | p-value<sup>7</sup> | r | p-value<sup>8</sup> |
| **231.0839**<sup>9</sup> | **0.89** | RP+ | Unknown | 58378 | 18.5 | 0.41 | 1.2 x 10<sup>-19</sup> | 4.4 x 10<sup>-16</sup> | 0.38 | 7.0 x 10<sup>-11</sup> | 0.40 | 6.3 x 10<sup>-18</sup> |
| 253.0925 | 0.93 | RP- | Unknown | 11140 | 13.2 | 0.39 | 2.6 x 10<sup>-18</sup> | 4.6 x 10<sup>-15</sup> | 0.32 | 3.2 x 10<sup>-8</sup> | -<sup>10</sup> | - |
| **203.0227**<sup>9</sup> | **2.78** | RP+ | 2-hydroxy-3-methylbutyric acid | 204079 | 14.8 | 0.26 | 1.9 x 10<sup>-8</sup> | 2.0 x 10<sup>-6</sup> | 0.24 | 5.3 x 10<sup>-5</sup> | 0.40 | 1.1 x 10<sup>-18</sup> |
| 217.9895 | 2.78 | RP+ | 2-hydroxy-3-methylbutyric acid | 36539 | 11.7 | 0.30 | 9.0 x 10<sup>-11</sup> | 2.1 x 10<sup>-8</sup> | 0.25 | 2.3 x 10<sup>-5</sup> | 0.38 | 2.4 x 10<sup>-16</sup> |
| 250.0134 | 2.78 | RP+ | 2-hydroxy-3-methylbutyric acid | 122838 | 12.5 | 0.28 | 9.0 x 10<sup>-10</sup> | 1.6 x 10<sup>-7</sup> | 0.27 | 8.2 x 10<sup>-6</sup> | 0.40 | 3.5 x 10<sup>-18</sup> |
| 221.0605 | 2.78 | RP+ | 2-hydroxy-3-methylbutyric acid | 56192 | 11.2 | 0.28 | 2.6 x 10<sup>-9</sup> | 3.2 x 10<sup>-7</sup> | 0.25 | 2.1 x 10<sup>-5</sup> | 0.39 | 1.9 x 10<sup>-17</sup> |
| 218.9958 | 2.78 | RP+ | 2-hydroxy-3-methylbutyric acid | 115590 | 11.7 | 0.28 | 1.3 x 10<sup>-9</sup> | 2.1 x 10<sup>-7</sup> | 0.26 | 1.8 x 10<sup>-5</sup> | 0.40 | 1.7 x 10<sup>-18</sup> |
| 235.0479 | 2.78 | RP+ | 2-hydroxy-3-methylbutyric acid | 34447 | 15.5 | 0.20 | 2.3 x 10<sup>-5</sup> | 1.0 x 10<sup>-3</sup> | 0.26 | 2.1 x 10<sup>-5</sup> | 0.38 | 4.2 x 10<sup>-16</sup> |
| 117.0559 | 2.78 | RP- | 2-hydroxy-3-methylbutyric acid | 211842 | 12.1 | 0.28 | 1.3 x 10<sup>-9</sup> | 2.2 x 10<sup>-7</sup> | 0.28 | 2.0 x 10<sup>-6</sup> | -<sup>10</sup> | - |
| 261.9788 | 2.78 | RP- | 2-hydroxy-3-methylbutyric acid | 15985 | 11.9 | 0.27 | 7.2 x 10<sup>-9</sup> | 8.3 x 10<sup>-7</sup> | 0.28 | 2.7 x 10<sup>-6</sup> | -<sup>10</sup> | - |

<sup>1</sup> Quality control samples within the discovery set;
<sup>2</sup> The analyses of features acquired in positive and negative modes used data from 451 and 452 participants, respectively, after the exclusion of outliers and samples with too many missing values;
<sup>3</sup> The analyses of features acquired in positive and negative modes used data from 271 and 277 participants, respectively, after the exclusion of outliers and samples with too many missing values;
<sup>4</sup> *m/z*= monoisotopic mass divided by the charge state values, as observed in the discovery set;
<sup>5</sup> Retention time;
<sup>6</sup> Q-values associated to False Discovery Rate (FDR) procedure to correct for multiple testing [64], alpha=0.05;
<sup>7</sup> Threshold for statistical significance corrected with Bonferroni method for multiple testing, equal to 0.0007463 (0.05/*f*<sub>1</sub>, with *f*<sub>1</sub>=67).
<sup>8</sup> Threshold for statistical significance corrected with Bonferroni method for multiple testing, equal to 0.007 (0.05/*f*<sub>3</sub>, with *f*<sub>3</sub>=7);
<sup>9</sup> Feature chosen for analysis of disease see Table 3;
<sup>10</sup> Feature not available in ATBC.

For subsequent analyses, the feature with the greatest chromatographic intenstity (i.e., main feature) for each metabolite was used (**Table 2**). The discriminatory accuracy for high versus low alcohol consumption in cross-validated models that included both 2-hydroxy-3-methylbutyric acid and the unknown compound was 75% (95% CI: 69-86%) (**Figure 3**). The test for non-linearity using restricted regression spline was borderline significant (p=0.06; **Figure S1**). All seven positive ionization mode features selected in the EPIC replication set were confirmed in the ATBC replication set (**Table 2**). In the ATBC replication set, the discriminatory accuracy for alcohol consumption (high vs. low consumers as defined in men in EPIC) of 2-hydroxy-3-methylbutyric acid and the unknown compound was 76% (95% CI: 68-84%), only 2% higher than the discriminatory accuracy observed for 2-hydroxy-3-methylbutyric acid alone (74%, 95% CI: 66-82%).

**Figure 3:**
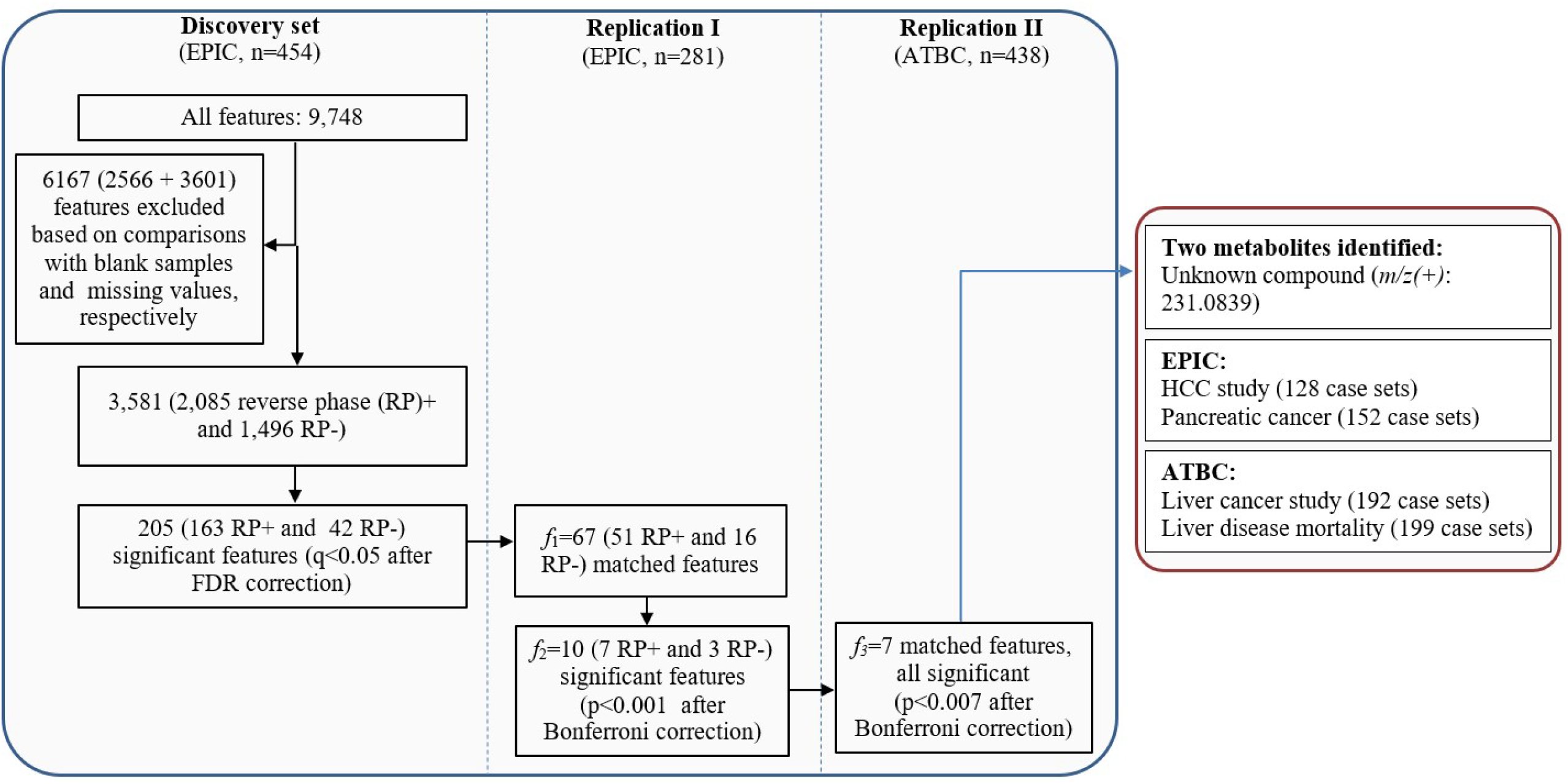
ROC curves with AUC for high alcohol intake, in the range 33.1-138.9 g/day in men (EPIC n=36; ATBC n=70) and 12.3-93.5 g/day in women (EPIC n=26), *vs*. low alcohol intake, in the range of >0.1-4.9 g/day in men (EPIC n=35; ATBC n=90) and >0.1-1.0 g/day in women (EPIC n=25), in **(A)** EPIC replication set and **(B)** ATBC replication set for the two identified biomarker candidates after cross-validation.

### Disease risk associations

In multivariable models (**Table 3**), 2-hydroxy-3-methylbutyric acid was associated with increased odds of HCC (OR_1-SD_=2.14: 1.39, 3.31) and pancreatic cancer (OR_1-SD_=1.65: 1.17, 2.32) in EPIC, as well as liver cancer (OR_1-SD_=2.00; 1.44, 2.77) and fatal liver disease (OR_1-SD_=2.16; 1.63, 2.86) in ATBC; the unknown candidate biomarker was associated with increased odds of liver cancer (OR_1-SD_=1.70; 95% CI: 1.29, 2.25) and liver disease mortality (OR=1.85; 95% CI: 1.39-2.46) in ATBC, but not with HCC or pancreatic cancer in EPIC. Alcohol intake was not associated with HCC (OR_1-SD_=0.78; 95% CI: 0.56, 1.09) or pancreatic cancer risk (OR_1-SD_=1.03: 0.77, 1.39) in EPIC, but was strongly associated with liver disease mortality (OR_1-SD_=2.19: 95% CI, 1.60, 2.98) in ATBC. The alcohol findings are in line with previously published EPIC and ATBC analyses [39-41].

**Table 3.**
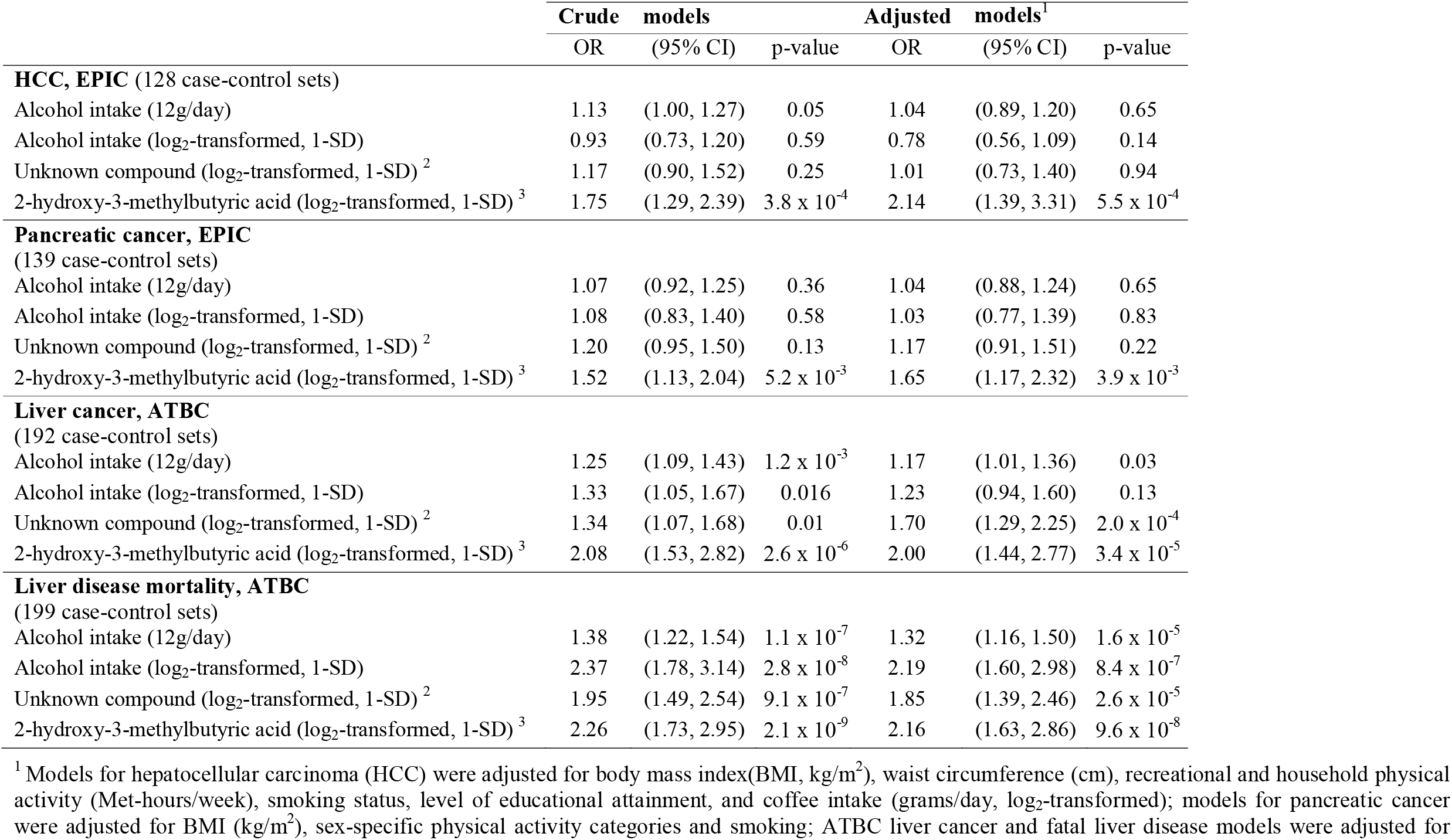
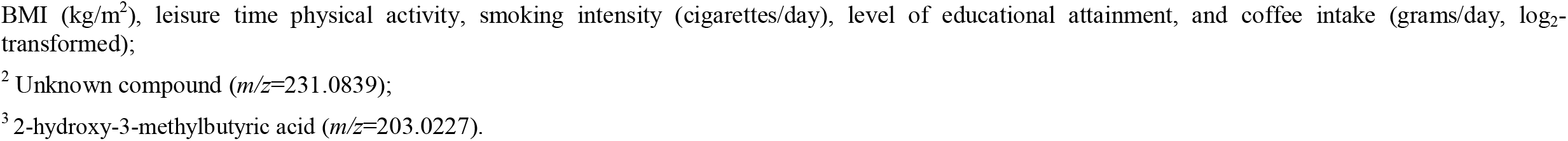
Crude and adjusted ^1^ odds ratios (OR, 95% CI) of self-reported alcohol intake (12 g/day) and the main features of the unknown compound and 2-hydroxy-3-methylbutyric acid (per 1-SD) with hepatocellular carcinoma (HCC; 129 case-control sets) and pancreatic cancer (152 case-control sets) in EPIC, and with liver cancer (194 case-control sets) and liver disease mortality (201 case-control sets) in ATBC

|  | **Crude models** |  |  | **Adjusted models<sup>1</sup>** |  |  |
| --- | --- | --- | --- | --- | --- | --- |
|  | OR | (95% CI) | p-value | OR | (95% CI) | p-value |
| **HCC, EPIC** (128 case-control sets) |  |  |  |  |  |  |
| Alcohol intake (12g/day) | 1.13 | (1.00, 1.27) | 0.05 | 1.04 | (0.89, 1.20) | 0.65 |
| Alcohol intake (log<sub>2</sub>-transformed, 1-SD) | 0.93 | (0.73, 1.20) | 0.59 | 0.78 | (0.56, 1.09) | 0.14 |
| Unknown compound (log<sub>2</sub>-transformed, 1-SD) <sup>2</sup> | 1.17 | (0.90, 1.52) | 0.25 | 1.01 | (0.73, 1.40) | 0.94 |
| 2-hydroxy-3-methylbutyric acid (log<sub>2</sub>-transformed, 1-SD) <sup>3</sup> | 1.75 | (1.29, 2.39) | 3.8 x 10<sup>-4</sup> | 2.14 | (1.39, 3.31) | 5.5 x 10<sup>-4</sup> |
| **Pancreatic cancer, EPIC** (139 case-control sets) |  |  |  |  |  |  |
| Alcohol intake (12g/day) | 1.07 | (0.92, 1.25) | 0.36 | 1.04 | (0.88, 1.24) | 0.65 |
| Alcohol intake (log<sub>2</sub>-transformed, 1-SD) | 1.08 | (0.83, 1.40) | 0.58 | 1.03 | (0.77, 1.39) | 0.83 |
| Unknown compound (log<sub>2</sub>-transformed, 1-SD) <sup>2</sup> | 1.20 | (0.95, 1.50) | 0.13 | 1.17 | (0.91, 1.51) | 0.22 |
| 2-hydroxy-3-methylbutyric acid (log<sub>2</sub>-transformed, 1-SD) <sup>3</sup> | 1.52 | (1.13, 2.04) | 5.2 x 10<sup>-3</sup> | 1.65 | (1.17, 2.32) | 3.9 x 10<sup>-3</sup> |
| **Liver cancer, ATBC** (192 case-control sets) |  |  |  |  |  |  |
| Alcohol intake (12g/day) | 1.25 | (1.09, 1.43) | 1.2 x 10<sup>-3</sup> | 1.17 | (1.01, 1.36) | 0.03 |
| Alcohol intake (log<sub>2</sub>-transformed, 1-SD) | 1.33 | (1.05, 1.67) | 0.016 | 1.23 | (0.94, 1.60) | 0.13 |
| Unknown compound (log<sub>2</sub>-transformed, 1-SD) <sup>2</sup> | 1.34 | (1.07, 1.68) | 0.01 | 1.70 | (1.29, 2.25) | 2.0 x 10<sup>-4</sup> |
| 2-hydroxy-3-methylbutyric acid (log<sub>2</sub>-transformed, 1-SD) <sup>3</sup> | 2.08 | (1.53, 2.82) | 2.6 x 10<sup>-6</sup> | 2.00 | (1.44, 2.77) | 3.4 x 10<sup>-5</sup> |
| **Liver disease mortality, ATBC** (199 case-control sets) |  |  |  |  |  |  |
| Alcohol intake (12g/day) | 1.38 | (1.22, 1.54) | 1.1 x 10<sup>-7</sup> | 1.32 | (1.16, 1.50) | 1.6 x 10<sup>-5</sup> |
| Alcohol intake (log<sub>2</sub>-transformed, 1-SD) | 2.37 | (1.78, 3.14) | 2.8 x 10<sup>-8</sup> | 2.19 | (1.60, 2.98) | 8.4 x 10<sup>-7</sup> |
| Unknown compound (log<sub>2</sub>-transformed, 1-SD) <sup>2</sup> | 1.95 | (1.49, 2.54) | 9.1 x 10<sup>-7</sup> | 1.85 | (1.39, 2.46) | 2.6 x 10<sup>-5</sup> |
| 2-hydroxy-3-methylbutyric acid (log<sub>2</sub>-transformed, 1-SD) <sup>3</sup> | 2.26 | (1.73, 2.95) | 2.1 x 10<sup>-9</sup> | 2.16 | (1.63, 2.86) | 9.6 x 10<sup>-8</sup> |

<sup>1</sup> Models for hepatocellular carcinoma (HCC) were adjusted for body mass index(BMI, kg/m<sup>2</sup>), waist circumference (cm), recreational and household physical activity (Met-hours/week), smoking status, level of educational attainment, and coffee intake (grams/day, log<sub>2</sub>-transformed); models for pancreatic cancer were adjusted for BMI (kg/m<sup>2</sup>), sex-specific physical activity categories and smoking; ATBC liver cancer and fatal liver disease models were adjusted for
BMI ( $\text{kg}/\text{m}^2$ ), leisure time physical activity, smoking intensity (cigarettes/day), level of educational attainment, and coffee intake (grams/day, $\log_2$ -transformed);
<sup>2</sup> Unknown compound ( $m/z=231.0839$ );
<sup>3</sup> 2-hydroxy-3-methylbutyric acid ( $m/z=203.0227$ ).

## DISCUSSION

Using untargeted metabolomics data from a discovery and two independent replication sets, we found two serum metabolites that were highly correlated with self-reported habitual alcohol intake. One compound was identified as 2-hydroxy-3-methylbutyric acid; the other remains unknown but is likely a combination of isomers of ethyl glucoside. Of note, ethyl-α-D-glucoside is a known constituent of some alcoholic beverages [38]. Both compounds could discriminate high alcohol consumers from low consumers in EPIC and ATBC despite marked differences in the study designs. Notably, 2-hydroxy-3-methylbutyric acid was strongly associated with HCC and pancreatic cancer risks in EPIC, and with liver cancer and fatal liver disease in ATBC. In contrast, self-reported alcohol intake was only consistently associated with risk of liver disease mortality in ATBC. Further research is needed to elucidate the potential metabolic cascade from alcohol drinking to 2-hydroxy-3-methylbutyric acid to disease. Additionally, studies measuring circulating concentrations of 2-hydroxy-3-methylbutyric acid rather than relative levels are now needed to compare across studies and improve risk estimation; this is especially important for diseases such as pancreatic cancer, for which the literature is suggestive [42] yet inconsistent [43].

Prior population-based studies have used a targeted or semi-targeted metabolomics approach to identify alcohol-specific metabolomic profiles of alcohol intake. Three studies, including one in EPIC, used targeted metabolomics, measuring 123 to 163 metabolites, to gain insight into metabolic pathways linking alcohol drinking to human health [44-46]; ten alcohol-metabolite associations were common to all three studies and included phosphatidylcholines (PCs), LysoPCs, acylcarnitines and sphingomyelins. Of note, PCs contribute to the formation of PEth in human tissues [47], which is a known biomarker of recent and heavy alcohol consumption used to diagnose alcohol abuse [48, 49]. A fourth targeted study used nuclear magnetic resonance to evaluate cross-sectional associations of 76 lipids, fatty acids, amino acids, ketone bodies and gluconeogenesis-related metabolites with alcohol consumption [50]. The endogenous metabolites identified by these targeted platforms did not overlap with the compounds most highly correlated with alcohol intake in our untargeted study, underscoring the breadth of the metabolome and discovery potential of untargeted metabolomics methods.

Metabolomics analyses that limit biomarker discovery to previously annotated compounds have also identified several alcohol-related biomarkers. For example, using prediagnostic serum samples from a nested breast cancer case-control study within a U.S. cohort, alcohol intake was associated with 16 of the 617 annotated metabolites, including 2-hydroxy-3-methylbutyric acid, 2,3-dihydroxyisovaleric acid (i.e., 2,3-hydroxy-3-methylbutyric acid), ethyl glucuronide and several endogenous metabolites related to androgen metabolism [51]. Other cross-sectional analyses, measuring hundreds of metabolites, also found associations of 2-hydroxy-3-methylbutyric acid, 2,3-dihydroxyisovaleric acid (i.e., 2,3-hydroxy-2-methylbutyric acid) and ethyl glucuronide with alcohol intake using prediagnostic serum [52, 53]. However, these studies lacked separate discovery and replication steps, and estimated correlations in cases and controls combined rather than in controls only. One study, which included discovery and replication sets, evaluated associations between alcohol intake and 356 known metabolites among African Americans [54] and found that alcohol was associated with five 2-hydroxybutyrate-related metabolites including 2-hydroxy-3-methylbutyric acid. A Japanese study of 107 metabolites in men identified positive associations between 2-hydroxybutyric acid and alcohol intake in a discovery and a replication set [55]. The production of 2-hydroxy-3-methylbutyric acid and other hydroxybutyric acid-related metabolites is linked to the rate of hepatic glutathione synthesis, which can increase considerably in response to oxidative stress or detoxification of xenobiotics in the liver [56]. A targeted metabolomics investigation in EPIC found evidence suggesting that glutathione metabolism is involved in the development of HCC [20]. Additionally, 2-hydroxy-3-methylbutyric acid is a product of branched-chain amino acid metabolism, which has been linked to alcohol drinking [55, 57].

To our knowledge, this study is unique in its untargeted metabolomics approach without preselected metabolites and its use of a discovery and two independent replication sets. By considering nearly 7,000 features, many of which are correlated, we greatly increased the number of potential candidates, but we also incurred a stronger penalisation for multiple testing. Consequently, our approach may have missed features that did not meet stringent thresholds for statistical significance. A strength of our approach was the use of large independent discovery and replications sets; although matching features by mass and retention time across sets may have resulted in the loss of relevant information. Other potential limitations relate to generalizability and measurement error. Circulating metabolite levels reflect environmental exposures as well as host and microbial metabolism [58-60], and identification of candidate biomarkers that are sufficiently specific to ethanol and generalizable to diverse populations is challenging. Measurement error, both systematic and random, is inherent to self-reported assessments [61-63], including alcohol intake, and likely biases association estimates in not only aetiological studies, but also in biomarker discovery studies. Despite our use of cutting-edge untargeted metabolomics methods, a robust study design, and an aetiological component to evaluate the associations of our candidate biomarkers with disease outcomes, we cannot dismiss the possibility that our findings were impacted by measurement error in self-reported alcohol intake.

In summary, we observed robust correlations between self-reported habitual alcohol intake and 2-hydroxy-3-methylbutyric acid and an unidentified compound in a discovery set and two independent replication sets of cancer-free participants. Associations for 2-hydroxy-3-methylbutyric acid with risk of HCC and pancreatic cancer in the EPIC study and with liver cancer in ATBC were stronger than those for either self-reported alcohol intake or the unidentified compound. In conclusion, 2-hydroxy-3-methylbutyric acid is a promising candidate biomarker for studying the relationship between habitual alcohol intake and health [51-54], but further research, preferably in the context a randomized-controlled trial, is needed to better characterize the relationship between 2-hydroxy-3-methylbutyric acid and alcohol at varying levels of intake.

## Data Availability

Access to EPIC data and biospecimens can be found at http://epic.iarc.fr/access/index.php

## Acknowledgements

EPIC Umeå investigators thank the Västerbotten Intervention Programme and the County Council of Västerbotten for providing data and samples and acknowledge the contribution from Biobank Sweden, supported by the Swedish Research Council (VR 2017-00650). We thank the National Institute for Public Health and the Environment (RIVM), Bilthoven, the Netherlands, for their contribution and ongoing support to the EPIC Study.

## Availability of data and materials

For information on how to submit an application for gaining access to EPIC data and/or biospecimens, please follow the instructions at http://epic.iarc.fr/access/index.php

