## Supplementary material for "Novel biomarkers of habitual alcohol intake and associations with risk of pancreatic and liver cancers and liver disease mortality": Supp. Material

**Table S1**. Correlations of alcohol intake with 205 (out of the remaining 3,581) features in the EPIC cross-sectional discovery dataset (n=454) that were statistically significant and carried forward for replication

| m/z | Retention time (min) | Method | r with alcohol | p-value | q-value* |
|---|---|---|---|---|---|
| 231.08388 | 0.892 | RP+ | 0.41 | 1.23E-19 | 4.39E-16 |
| 253.09252 | 0.934 | RP- | 0.39 | 2.58E-18 | 4.62E-15 |
| 226.12958 | 1.426 | RP+ | 0.39 | 6.73E-18 | 8.04E-15 |
| 308.02038 | 0.892 | RP+ | 0.35 | 8.68E-15 | 7.77E-12 |
| 231.08408 | 1.411 | RP+ | 0.34 | 6.35E-14 | 4.55E-11 |
| 788.54092 | 8.517 | RP- | -0.33 | 5.77E-13 | 3.44E-10 |
| 290.15908 | 1.947 | RP+ | 0.33 | 1.38E-12 | 7.08E-10 |
| 526.31512 | 6.887 | RP- | -0.32 | 2.08E-12 | 9.30E-10 |
| 594.30112 | 6.886 | RP- | -0.32 | 4.14E-12 | 1.65E-09 |
| 783.06838 | 9.111 | RP+ | 0.31 | 8.85E-12 | 3.17E-09 |
| 779.55778 | 9.118 | RP+ | 0.31 | 2.68E-11 | 8.73E-09 |
| 391.78018 | 9.131 | RP+ | 0.3 | 6.85E-11 | 2.04E-08 |
| 772.07498 | 9.122 | RP+ | 0.3 | 7.95E-11 | 2.10E-08 |
| 780.06058 | 9.119 | RP+ | 0.3 | 8.22E-11 | 2.10E-08 |
| 217.98948 | 2.777 | RP+ | 0.3 | 8.96E-11 | 2.14E-08 |
| 813.55718 | 9.116 | RP+ | 0.29 | 2.19E-10 | 4.89E-08 |
| 220.99658 | 2.778 | RP+ | 0.29 | 5.49E-10 | 1.16E-07 |
| 71.05012 | 2.784 | RP- | 0.28 | 8.25E-10 | 1.61E-07 |
| 760.58308 | 9.108 | RP+ | 0.28 | 8.56E-10 | 1.61E-07 |
| 250.01338 | 2.777 | RP+ | 0.28 | 9.01E-10 | 1.61E-07 |
| 218.99578 | 2.777 | RP+ | 0.28 | 1.25E-09 | 2.14E-07 |
| 117.05592 | 2.782 | RP- | 0.28 | 1.33E-09 | 2.16E-07 |
| 482.32258 | 6.889 | RP+ | -0.28 | 1.39E-09 | 2.16E-07 |
| 222.99018 | 2.778 | RP+ | 0.28 | 1.98E-09 | 2.85E-07 |
| 399.77128 | 9.127 | RP+ | 0.28 | 1.99E-09 | 2.85E-07 |
| 788.05228 | 9.119 | RP+ | 0.28 | 2.09E-09 | 2.87E-07 |
| 407.75808 | 9.128 | RP+ | 0.28 | 2.35E-09 | 3.12E-07 |
| 129.01912 | 1.374 | RP- | 0.28 | 2.51E-09 | 3.21E-07 |
| 221.06048 | 2.778 | RP+ | 0.28 | 2.62E-09 | 3.23E-07 |
| 400.27318 | 9.127 | RP+ | 0.27 | 3.63E-09 | 4.34E-07 |
| 261.97882 | 2.784 | RP- | 0.27 | 7.19E-09 | 8.30E-07 |
| 251.94542 | 2.784 | RP- | 0.27 | 8.06E-09 | 8.77E-07 |
| 744.55208 | 8.513 | RP+ | -0.27 | 8.08E-09 | 8.77E-07 |
| 782.57038 | 9.1 | RP+ | 0.26 | 1.57E-08 | 1.66E-06 |
| 203.02268 | 2.778 | RP+ | 0.26 | 1.92E-08 | 1.96E-06 |
| 175.05718 | 0.698 | RP+ | 0.26 | 2.14E-08 | 2.13E-06 |
| 392.28408 | 9.132 | RP+ | 0.26 | 2.20E-08 | 2.13E-06 |
| 378.90542 | 2.784 | RP- | 0.26 | 2.29E-08 | 2.16E-06 |
| 408.25738 | 9.128 | RP+ | 0.26 | 2.94E-08 | 2.70E-06 |
| 126.05448 | 0.887 | RP+ | 0.25 | 5.86E-08 | 5.25E-06 |
| 782.57178 | 8.673 | RP+ | 0.24 | 1.39E-07 | 1.21E-05 |
| 787.54688 | 9.118 | RP+ | 0.24 | 1.95E-07 | 1.66E-05 |
| 393.74338 | 8.605 | RP+ | 0.24 | 2.17E-07 | 1.79E-05 |
| 732.55278 | 8.606 | RP+ | 0.24 | 2.20E-07 | 1.79E-05 |
| 403.27468 | 8.669 | RP+ | 0.24 | 2.52E-07 | 2.00E-05 |
| 261.14018 | 6.885 | RP+ | -0.24 | 2.76E-07 | 2.15E-05 |
| 329.03968 | 3.214 | RP+ | 0.23 | 6.90E-07 | 5.26E-05 |
| 776.54462 | 8.61 | RP- | 0.23 | 7.29E-07 | 5.44E-05 |
| 402.77348 | 8.67 | RP+ | 0.23 | 7.72E-07 | 5.64E-05 |
| 326.03218 | 3.213 | RP+ | 0.23 | 9.72E-07 | 6.96E-05 |
| 802.04698 | 8.668 | RP+ | 0.23 | 1.20E-06 | 8.19E-05 |

| | | | | | |
|---|---|---|---|---|---|
| 217.07028 | 3.214 | RP+ | 0.23 | 1.21E-06 | 8.19E-05 |
| 463.74368 | 8.786 | RP+ | -0.23 | 1.21E-06 | 8.19E-05 |
| 151.06032 | 0.708 | RP- | 0.23 | 1.28E-06 | 8.52E-05 |
| 211.14568 | 3.878 | RP+ | 0.22 | 1.59E-06 | 1.04E-04 |
| 816.57122 | 9.008 | RP- | -0.22 | 1.68E-06 | 1.07E-04 |
| 197.06612 | 0.708 | RP- | 0.22 | 2.15E-06 | 1.35E-04 |
| 510.35388 | 7.136 | RP+ | -0.22 | 2.42E-06 | 1.49E-04 |
| 554.34732 | 7.137 | RP- | -0.22 | 2.45E-06 | 1.49E-04 |
| 377.76478 | 8.606 | RP+ | 0.22 | 2.52E-06 | 1.50E-04 |
| 800.03138 | 8.425 | RP+ | 0.22 | 3.01E-06 | 1.77E-04 |
| 195.08898 | 3.213 | RP+ | 0.22 | 3.11E-06 | 1.80E-04 |
| 622.33162 | 7.132 | RP- | -0.21 | 4.74E-06 | 2.70E-04 |
| 801.54538 | 8.668 | RP+ | 0.21 | 5.12E-06 | 2.87E-04 |
| 782.05728 | 8.703 | RP+ | 0.21 | 5.31E-06 | 2.92E-04 |
| 464.74798 | 9.11 | RP+ | 0.21 | 5.95E-06 | 3.23E-04 |
| 385.75488 | 8.605 | RP+ | 0.21 | 6.07E-06 | 3.24E-04 |
| 210.04858 | 9.053 | RP+ | -0.21 | 6.69E-06 | 3.53E-04 |
| 275.15658 | 7.133 | RP+ | -0.21 | 7.30E-06 | 3.79E-04 |
| 817.06528 | 9.131 | RP+ | 0.21 | 8.48E-06 | 4.34E-04 |
| 789.03828 | 8.602 | RP+ | 0.21 | 9.90E-06 | 4.99E-04 |
| 386.25628 | 8.604 | RP+ | 0.21 | 1.11E-05 | 5.54E-04 |
| 194.08008 | 3.218 | RP+ | 0.2 | 1.30E-05 | 6.39E-04 |
| 844.53968 | 9.121 | RP+ | 0.2 | 1.37E-05 | 6.64E-04 |
| 934.53112 | 9.127 | RP- | 0.2 | 1.48E-05 | 7.07E-04 |
| 447.78298 | 7.157 | RP+ | -0.2 | 1.87E-05 | 8.82E-04 |
| 311.06468 | 3.214 | RP+ | 0.2 | 2.03E-05 | 9.33E-04 |
| 410.25698 | 8.418 | RP+ | 0.2 | 2.03E-05 | 9.33E-04 |
| 872.56142 | 9.126 | RP- | 0.2 | 2.14E-05 | 9.68E-04 |
| 235.04788 | 2.778 | RP+ | 0.2 | 2.34E-05 | 1.04E-03 |
| 405.77078 | 8.988 | RP+ | -0.2 | 2.36E-05 | 1.04E-03 |
| 504.30458 | 6.888 | RP+ | -0.2 | 2.72E-05 | 1.19E-03 |
| 804.57622 | 9.126 | RP- | 0.19 | 3.22E-05 | 1.39E-03 |
| 409.75498 | 8.418 | RP+ | 0.19 | 3.44E-05 | 1.46E-03 |
| 181.07238 | 2.824 | RP+ | 0.19 | 3.46E-05 | 1.46E-03 |
| 279.04088 | 3.216 | RP+ | 0.19 | 3.55E-05 | 1.48E-03 |
| 734.56828 | 9.025 | RP+ | 0.19 | 3.64E-05 | 1.50E-03 |
| 410.76218 | 8.668 | RP+ | 0.19 | 3.87E-05 | 1.58E-03 |
| 166.07258 | 0.901 | RP+ | 0.19 | 4.15E-05 | 1.64E-03 |
| 411.26398 | 8.667 | RP+ | 0.19 | 4.19E-05 | 1.64E-03 |
| 439.79508 | 7.153 | RP+ | -0.19 | 4.19E-05 | 1.64E-03 |
| 230.09588 | 0.751 | RP+ | 0.19 | 4.22E-05 | 1.64E-03 |
| 809.53008 | 8.667 | RP+ | 0.19 | 4.34E-05 | 1.67E-03 |
| 826.55792 | 8.673 | RP- | 0.19 | 4.39E-05 | 1.67E-03 |
| 802.53878 | 8.42 | RP+ | 0.19 | 5.02E-05 | 1.89E-03 |
| 176.12778 | 0.761 | RP+ | 0.19 | 5.95E-05 | 2.22E-03 |
| 502.29158 | 6.896 | RP+ | 0.19 | 6.18E-05 | 2.28E-03 |
| 790.04748 | 8.699 | RP+ | 0.19 | 6.59E-05 | 2.41E-03 |
| 159.06238 | 0.767 | RP+ | 0.19 | 6.87E-05 | 2.48E-03 |
| 524.27578 | 6.894 | RP+ | 0.19 | 6.97E-05 | 2.49E-03 |
| 633.48278 | 7.44 | RP+ | -0.18 | 7.75E-05 | 2.75E-03 |
| 780.55678 | 8.421 | RP+ | 0.18 | 8.35E-05 | 2.91E-03 |
| 799.52948 | 8.422 | RP+ | 0.18 | 8.37E-05 | 2.91E-03 |
| 940.54482 | 9.126 | RP- | 0.18 | 8.52E-05 | 2.92E-03 |
| 417.74148 | 8.417 | RP+ | 0.18 | 8.56E-05 | 2.92E-03 |
| 806.56958 | 8.602 | RP+ | 0.18 | 8.78E-05 | 2.96E-03 |
| 286.14508 | 5.992 | RP+ | 0.18 | 8.83E-05 | 2.96E-03 |

| | | | | | |
|---|---|---|---|---|---|
| 384.27268 | 8.586 | RP+ | 0.18 | 9.27E-05 | 3.08E-03 |
| 632.98048 | 7.439 | RP+ | -0.18 | 1.01E-04 | 3.30E-03 |
| 778.53508 | 8.421 | RP+ | 0.18 | 1.01E-04 | 3.30E-03 |
| 647.55788 | 8.634 | RP+ | -0.18 | 1.07E-04 | 3.44E-03 |
| 844.53022 | 8.611 | RP- | 0.18 | 1.12E-04 | 3.58E-03 |
| 269.22698 | 7.225 | RP+ | 0.18 | 1.14E-04 | 3.62E-03 |
| 950.50352 | 9.127 | RP- | 0.18 | 1.24E-04 | 3.90E-03 |
| 626.45838 | 7.432 | RP+ | 0.18 | 1.26E-04 | 3.92E-03 |
| 789.54698 | 8.699 | RP+ | 0.18 | 1.36E-04 | 4.19E-03 |
| 418.74918 | 8.668 | RP+ | 0.18 | 1.40E-04 | 4.29E-03 |
| 794.06168 | 8.67 | RP+ | 0.18 | 1.45E-04 | 4.41E-03 |
| 956.51372 | 8.675 | RP- | 0.18 | 1.47E-04 | 4.42E-03 |
| 766.57428 | 8.902 | RP+ | 0.18 | 1.54E-04 | 4.61E-03 |
| 526.29288 | 6.889 | RP+ | 0.18 | 1.67E-04 | 4.93E-03 |
| 828.55188 | 8.456 | RP+ | 0.17 | 2.03E-04 | 5.97E-03 |
| 850.55648 | 9.114 | RP+ | 0.17 | 2.13E-04 | 6.16E-03 |
| 418.24448 | 8.418 | RP+ | 0.17 | 2.15E-04 | 6.16E-03 |
| 401.76518 | 8.419 | RP+ | 0.17 | 2.15E-04 | 6.16E-03 |
| 308.12608 | 5.991 | RP+ | 0.17 | 2.21E-04 | 6.28E-03 |
| 960.51942 | 8.419 | RP- | 0.17 | 2.33E-04 | 6.56E-03 |
| 419.25218 | 8.667 | RP+ | 0.17 | 2.55E-04 | 7.13E-03 |
| 824.54732 | 8.418 | RP- | 0.17 | 2.65E-04 | 7.36E-03 |
| 126.05428 | 2.817 | RP+ | -0.17 | 2.81E-04 | 7.74E-03 |
| 450.26162 | 6.81 | RP- | 0.17 | 3.12E-04 | 8.54E-03 |
| 592.26372 | 6.887 | RP- | 0.17 | 3.49E-04 | 9.46E-03 |
| 552.36662 | 7.236 | RP- | -0.17 | 3.62E-04 | 9.74E-03 |
| 124.04012 | 2.808 | RP- | -0.17 | 3.65E-04 | 9.76E-03 |
| 292.15008 | 6.905 | RP+ | 0.17 | 3.96E-04 | 1.05E-02 |
| 234.00418 | 1.781 | RP+ | 0.16 | 4.41E-04 | 1.15E-02 |
| 912.50532 | 8.668 | RP- | 0.16 | 4.42E-04 | 1.15E-02 |
| 106.04978 | 0.659 | RP+ | -0.16 | 4.62E-04 | 1.20E-02 |
| 892.53272 | 8.419 | RP- | 0.16 | 4.88E-04 | 1.26E-02 |
| 219.00882 | 1.787 | RP- | 0.16 | 5.29E-04 | 1.35E-02 |
| 762.98258 | 7.02 | RP+ | 0.16 | 5.59E-04 | 1.42E-02 |
| 312.01568 | 2.709 | RP+ | 0.16 | 5.78E-04 | 1.45E-02 |
| 826.04648 | 8.6 | RP+ | 0.16 | 5.79E-04 | 1.45E-02 |
| 788.61698 | 9.825 | RP+ | 0.16 | 6.98E-04 | 1.74E-02 |
| 762.50662 | 8.516 | RP- | 0.16 | 7.51E-04 | 1.85E-02 |
| 604.93898 | 7.179 | RP+ | -0.16 | 7.55E-04 | 1.85E-02 |
| 603.94768 | 7.186 | RP+ | -0.16 | 7.60E-04 | 1.85E-02 |
| 128.03148 | 0.651 | RP+ | -0.16 | 7.69E-04 | 1.86E-02 |
| 535.80808 | 7.065 | RP+ | 0.16 | 7.73E-04 | 1.86E-02 |
| 137.06968 | 0.621 | RP+ | 0.16 | 7.84E-04 | 1.87E-02 |
| 266.12548 | 0.898 | RP+ | -0.16 | 8.02E-04 | 1.89E-02 |
| 459.08328 | 3.209 | RP+ | 0.16 | 8.04E-04 | 1.89E-02 |
| 233.05738 | 1.78 | RP+ | 0.16 | 8.54E-04 | 2.00E-02 |
| 563.81618 | 6.907 | RP+ | 0.16 | 8.70E-04 | 2.02E-02 |
| 413.78638 | 9.827 | RP+ | 0.16 | 8.93E-04 | 2.06E-02 |
| 790.56618 | 9.05 | RP+ | 0.16 | 9.22E-04 | 2.12E-02 |
| 524.27592 | 6.887 | RP- | 0.15 | 9.57E-04 | 2.18E-02 |
| 484.73688 | 8.899 | RP+ | -0.15 | 9.78E-04 | 2.22E-02 |
| 402.76638 | 8.907 | RP+ | 0.15 | 9.94E-04 | 2.24E-02 |
| 452.27688 | 6.81 | RP+ | 0.15 | 1.01E-03 | 2.25E-02 |
| 476.74978 | 8.903 | RP+ | -0.15 | 1.02E-03 | 2.28E-02 |
| 814.04538 | 8.635 | RP+ | 0.15 | 1.04E-03 | 2.30E-02 |
| 763.48448 | 7.019 | RP+ | 0.15 | 1.06E-03 | 2.32E-02 |

| | | | | | |
|---|---|---|---|---|---|
| 471.72928 | 8.786 | RP+ | -0.15 | 1.21E-03 | 2.64E-02 |
| 972.48592 | 8.673 | RP- | 0.15 | 1.23E-03 | 2.67E-02 |
| 222.01958 | 1.78 | RP+ | 0.15 | 1.34E-03 | 2.89E-02 |
| 500.27838 | 6.764 | RP+ | 0.15 | 1.36E-03 | 2.91E-02 |
| 619.52708 | 10.833 | RP+ | 0.15 | 1.40E-03 | 2.98E-02 |
| 894.54722 | 8.673 | RP- | 0.15 | 1.42E-03 | 3.01E-02 |
| 211.14488 | 3.771 | RP+ | 0.15 | 1.46E-03 | 3.07E-02 |
| 202.08638 | 2.658 | RP+ | 0.15 | 1.48E-03 | 3.09E-02 |
| 424.79498 | 7.147 | RP+ | 0.15 | 1.53E-03 | 3.17E-02 |
| 179.05672 | 2.823 | RP- | 0.15 | 1.53E-03 | 3.17E-02 |
| 991.67548 | 7.017 | RP+ | 0.15 | 1.66E-03 | 3.42E-02 |
| 421.77178 | 9.825 | RP+ | 0.15 | 1.68E-03 | 3.44E-02 |
| 262.01358 | 1.78 | RP+ | 0.15 | 1.73E-03 | 3.52E-02 |
| 791.54018 | 8.415 | RP+ | 0.15 | 1.83E-03 | 3.70E-02 |
| 204.97948 | 1.78 | RP+ | 0.15 | 1.87E-03 | 3.76E-02 |
| 568.26442 | 6.893 | RP- | 0.15 | 1.92E-03 | 3.83E-02 |
| 516.30478 | 6.838 | RP+ | 0.15 | 1.93E-03 | 3.83E-02 |
| 221.03198 | 1.781 | RP+ | 0.15 | 1.94E-03 | 3.83E-02 |
| 206.98068 | 1.78 | RP+ | 0.15 | 1.95E-03 | 3.83E-02 |
| 297.94472 | 1.787 | RP- | 0.15 | 1.97E-03 | 3.85E-02 |
| 813.54488 | 8.634 | RP+ | 0.15 | 1.99E-03 | 3.88E-02 |
| 566.31928 | 6.899 | RP+ | 0.14 | 2.05E-03 | 3.94E-02 |
| 792.57528 | 9.022 | RP+ | 0.14 | 2.06E-03 | 3.94E-02 |
| 422.76228 | 8.454 | RP+ | 0.14 | 2.07E-03 | 3.94E-02 |
| 797.53018 | 8.699 | RP+ | 0.14 | 2.07E-03 | 3.94E-02 |
| 575.31108 | 6.9 | RP+ | 0.14 | 2.10E-03 | 3.99E-02 |
| 482.72148 | 8.416 | RP+ | 0.14 | 2.11E-03 | 3.99E-02 |
| 542.32308 | 6.774 | RP+ | 0.14 | 2.17E-03 | 4.07E-02 |
| 414.28648 | 9.827 | RP+ | 0.14 | 2.20E-03 | 4.10E-02 |
| 788.55798 | 8.906 | RP+ | 0.14 | 2.26E-03 | 4.20E-02 |
| 962.53462 | 8.671 | RP- | 0.14 | 2.29E-03 | 4.22E-02 |
| 423.26348 | 8.453 | RP+ | 0.14 | 2.32E-03 | 4.26E-02 |
| 300.28988 | 5.874 | RP+ | -0.14 | 2.35E-03 | 4.29E-02 |
| 203.97298 | 1.78 | RP+ | 0.14 | 2.46E-03 | 4.46E-02 |
| 810.03638 | 8.668 | RP+ | 0.14 | 2.50E-03 | 4.53E-02 |
| 58.06518 | 0.706 | RP+ | 0.14 | 2.53E-03 | 4.56E-02 |
| 415.27418 | 8.599 | RP+ | 0.14 | 2.58E-03 | 4.62E-02 |
| 821.52958 | 8.638 | RP+ | 0.14 | 2.66E-03 | 4.73E-02 |
| 770.46438 | 7.019 | RP+ | 0.14 | 2.67E-03 | 4.73E-02 |
| 500.27582 | 6.895 | RP- | 0.14 | 2.75E-03 | 4.85E-02 |
| 209.15318 | 6.311 | RP+ | 0.14 | 2.77E-03 | 4.85E-02 |
| 268.14128 | 0.906 | RP+ | -0.14 | 2.78E-03 | 4.86E-02 |
| * FDR corrected p-values | | | | | |

**Table S2.** Adjusted odds ratio[1] (OR, 95% CI) of HCC (129 case-control sets) and pancreatic cancer (152 case-control sets) in EPIC, and OR of liver cancer (194 case-control sets) and liver disease mortality (201 case-control sets) in ATBC associated to self-reported alcohol intake (12 g/day), the unknown compound and 2-hydroxy-3-methylbutyric acid (per 1-SD) excluding non-drinkers of alcohol (i.e. alcohol intake >0.1g/d).

| | **Adjusted models[1]** | | |
|---|---|---|---|
| | OR | (95% CI) | p-value |
| **HCC, EPIC** (129 case sets) | | | |
| Alcohol intake (12g/day) | 1.18 | (0.94, 1.47) | 0.15 |
| Alcohol intake ($log_2$-transformed, 1-SD) | 1.14 | (0.70, 1.87) | 0.60 |
| Unknown compound ($log_2$-transformed, 1-SD) | 1.33 | (0.85, 2.09) | 0.22 |
| 2-hydroxy-3-methylbutyric acid ($log_2$-transformed, 1-SD) | 2.30 | (1.32, 3.99) | 0.003 |
| **Pancreatic cancer, EPIC** (152 case sets) | | | |
| Alcohol intake (12g/day) | 1.00 | (0.79, 1.26) | 0.98 |
| Alcohol intake ($log_2$-transformed, 1-SD) | 0.90 | (0.58, 1.39) | 0.63 |
| Unknown compound ($log_2$-transformed, 1-SD) | 1.11 | (0.80, 1.53) | 0.52 |
| 2-hydroxy-3-methylbutyric acid ($log_2$-transformed, 1-SD) | 1.59 | (1.03, 2.44) | 0.04 |
| **Liver cancer, ATBC** (159 case-sets) | | | |
| Alcohol intake (12 g/day) | 1.17 | (0.99, 1.37) | 0.06 |
| Alcohol intake ($log_2$-transformed, 1-SD) | 1.35 | (1.00, 1.82) | 0.05 |
| Unknown compound ($log_2$-transformed, 1-SD) | 1.87 | (1.36, 2.57) | 0.0001 |
| 2-hydroxy-3-methylbutyric acid ($log_2$-transformed, 1-SD) | 1.79 | (1.26, 2.55) | 0.001 |
| **Liver disease mortality, ATBC** (176 case-sets) | | | |
| Alcohol intake (12 g/day) | 1.28 | (1.13, 1.45) | 0.0001 |
| Alcohol intake ($log_2$-transformed, 1-SD) | 1.98 | (1.46, 2.69) | $1.1 \times 10^{-5}$ |
| Unknown compound ($log_2$-transformed, 1-SD) | 1.85 | (1.39, 2.46) | $2.6 \times 10^{-5}$ |
| 2-hydroxy-3-methylbutyric acid ($log_2$-transformed, 1-SD) | 2.00 | (1.50, 2.67) | $2.7 \times 10^{-6}$ |

[1]Models for hepatocellular carcinoma (HCC) were adjusted for body mass index(BMI, kg/$m^2$), waist circumference (cm), recreational and household physical activity (Met-hours/week), smoking status, level of educational attainment, and coffee intake (grams/day, $log_2$-transformed); models for pancreatic cancer were adjusted for BMI (kg/$m^2$), sex-specific physical activity categories and smoking; ATBC liver cancer and fatal liver disease models were adjusted for BMI (kg/$m^2$), leisure time physical activity, smoking intensity (cigarettes/day), level of educational attainment, and coffee intake (grams/day, $log_2$-transformed).

**Figure S1.** Non-linear association between standardized residuals of 2-hydroxy-3-methylbutyric acid and alcohol intake with cubic regression splines (with 5 knots and p-value against the null hypothesis of linearity equal to 0.06).

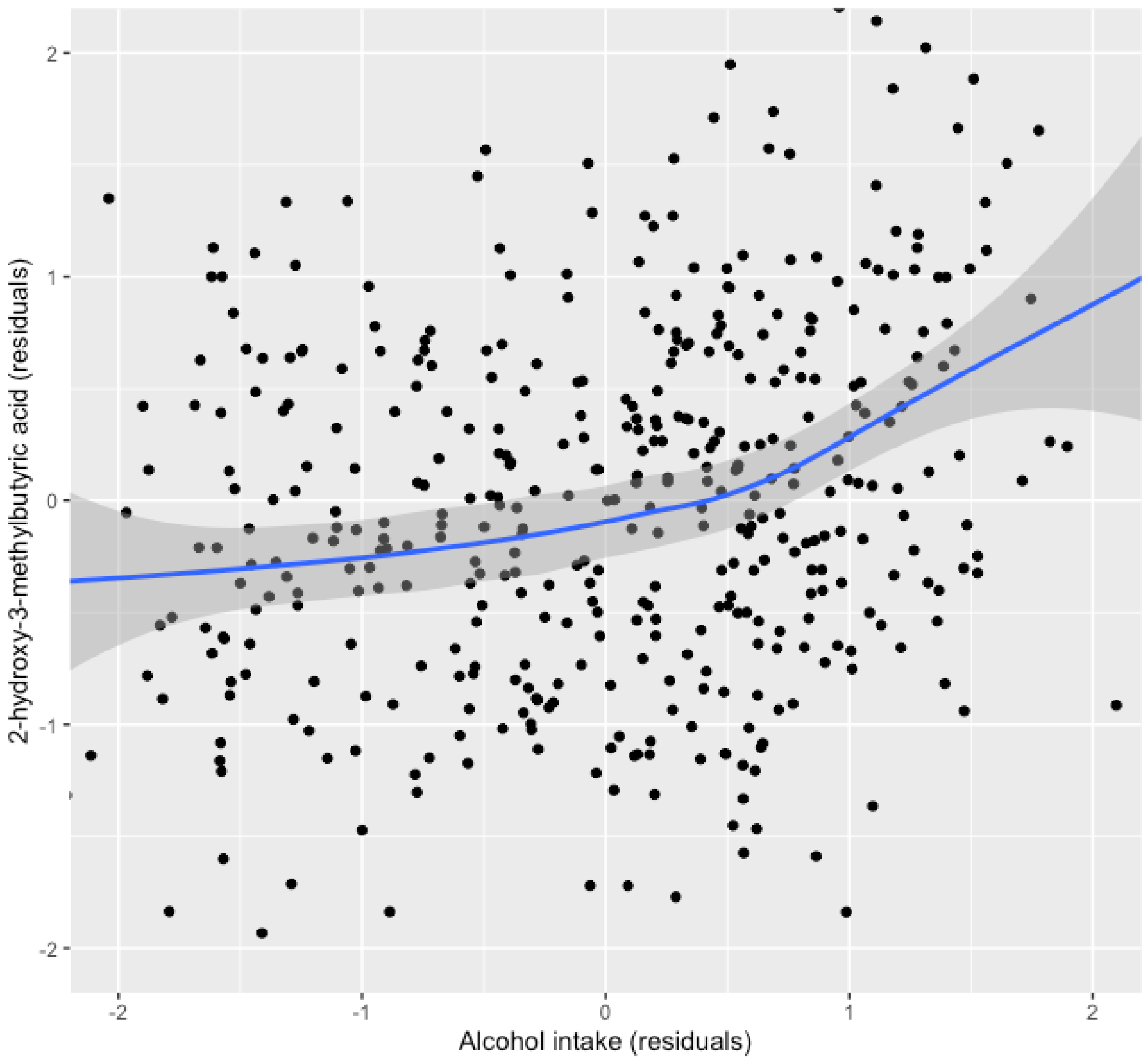

**Supplemental Methods: methods for sample analysis, data preprocessing, and feature identification**

*Sample preparation*

Samples were prepared by mixing 20, 25 or 30 µL of serum depending on study (cross sectional 20 µL, liver and pancreatic cancer 25 µL, ATBC 30 µL) with 200 µL of acetonitrile and vacuum filtering the precipitate with 0.2 µm Captiva ND plates (Agilent Technologies, Santa Clara, CA, USA). The filtrate was collected into a polypropylene well plate that was sealed (Rapid EPS, BioChromato, Fujisawa, Japan) and analyzed either immediately (cross sectional RP+, pancreatic cancer RP+, ATBC), after storing at -80 °C (liver cancer), or after drying measured aliquots and storing them at +4 °C (cross sectional RP-) or -80 °C (pancreatic cancer RP-) and reconstituting in their original volumes before analysis by adding 50% ACN. Quality control (QC) samples were prepared from a sample pool that was made by mixing small aliquots of at least 80 random samples. Blank samples were prepared in an identical manner, only leaving the serum out of the process. Each plate included four individually prepared QC samples and at least one blank.

*Sample analysis*

Samples were analyzed in positive and negative ionization mode for each of the three EPIC studies. The samples from the ATBC study were analyzed in positive ionization mode only. All samples were analyzed at the International Agency for Research on Cancer (IARC) as uninterrupted analytical runs for each ionization mode, except for the ATBC study that was analyzed as two consecutive batches of 511 and 513 samples with matched case-control sets placed consecutively but in random order. A UHPLC-QTOF-MS system was used that consisted of a 1290 Binary LC system, and a 6550 QTOF mass spectrometer with Jet Stream electrospray ionization source (Agilent Technologies). Samples were kept at 4°C and 2 µL was injected. An ACQUITY UHPLC HSS T3 column (2.1 × 100mm, 1.8 µm) was used at 45 °C and the mobile phase consisted of ultrapure water and LC-MS grade methanol, both with 0.05 % (v/v) of formic acid. The gradient profile was as follows: 0–6 min: 5% to 100% methanol, 6–10.5 min: 100% methanol, 10.5–13 min: 5% methanol. The flow rate was 0.4 ml/min.

Mass spectrometer drying gas temperature was 175°C and flow 12 L/min, with capillary, nozzle, and fragmentor voltages of 3500 V, 300 V, and 175 V, respectively. The sheath gas temperature was 350°C and flow 11 L/min, and nebulizer pressure 45 psi. MS/MS analyses were performed with an isolation width of 1.3 Da and collision energies of 10V, 20V, and 40 V. Continuous mass axis calibration was employed using Lock mass ions *m/z* 121.05087 and 922.00980 (positive) and *m/z* 112.98559 + 966.00073 (negative). Data was acquired in centroid format using an extended dynamic range mode, and acquisition rate of 1.67 Hz, over the mass range of 50-1000 Da (MassHunter Acquisition B.05.01, Agilent Technologies). The analytical run was initiated with priming injections of a QC sample to achieve stable instrument response, followed by a blank sample and the study samples, with one QC sample after every 10-12 injections. Quality assessment of the raw data before any transformation or adjustment was based on monitoring of known target metabolites in the QC samples. Intra-batch coefficient of variation (CV) of four representative metabolites hippuric acid, indole lactic acid, phenylalanine and tryptophan ranged from 3.3% to 16.5% (n=23−45, depending on batch size), with retention times within 0.15 min across all the datasets. After identification of the candidate biomarkers, their CV in the discovery dataset QC samples was calculated and reported in Table 2 of the main paper.

*Data pre-processing*

Pre-processing was performed using Agilent recursive feature finding workflow using MassHunter Qualitative Analysis B.06.00, DA Reprocessor, and Mass Profiler Professional 12.1 software, except for the pancreatic cancer study, for which the targeted feature extraction part of the workflow was performed with Agilent ProFinder 08.00. Data from each analytical batch was processed separately. The initial processing was performed using molecular feature extraction (MFE) algorithm for small molecules using data of the study samples only. Mass peak height threshold of 1500 and 300 counts was used for positive and negative mode data, with thresholds for chromatographic peaks at 10000 and 2000 counts, respectively. Spacing tolerance for possible isotope peaks was 0.0025 m/z plus 7 ppm, with a grouping model for common organic molecules. Note herein, we refer to "feature" as a chromatographic peak found by the preprocessing algorithm, while "compound" or "metabolite" refer to a confirmed molecule that can consist of one or more features that may be parent ions, adducts or fragments. Chromatographic peak areas were used as a measure of intensity. Feature alignment across the samples was performed with retention time and mass windows of 0.07 min and 15 ppm + 2 mDa, respectively. A target list for the recursive extraction was then created by including the features found in at least 2% of the samples (in at least 10 of the samples for pancreatic cancer). Recursive feature extraction of data from study samples, QCs and blanks was then performed using ±10 ppm and ±0.035 min match tolerances. Throughout the process, ion species were limited to $[M+H]^+$ and $[M-H]^-$. Resulting features were finally aligned using the same parameters as above, and peak areas were used as a measurement of intensity. The two datasets from the ATBC study were aligned into a single dataset at the end of the recursive process using retention time and mass windows of 0.08 min and 15 ppm + 2 mDa, respectively. For quality control, Find by Formula algorithm in Qualitative Analysis B.06.00 was used with metabolite formulas as targets, using same settings for chromatogram extraction and matching as above.

*Matching features across datasets*

Alcohol-associated features found in the discovery dataset were matched to features in the replication datasets of the same ionization mode based on closeness of the mass and retention time, by using ±15 ppm and ±0.2 min matching tolerance. In case multiple matching features were found in the replication dataset, matching was confirmed by visually comparing the chromatograms of the feature in a QC sample from both studies.

*Identification of molecular features*

Features were clustered by retention time, mass, and intensity correlation across the samples to help in finding features originating from the same compound, by using in-house developed software. The data used for identification was from the cross-sectional study. The m/z values were searched against the Human Metabolite Database (HMDB)[1] and METLIN metabolite database[2] using ions $[M+H]^+$, $[M+Na]^+$, $[M-H]^-$, $[M+FA-H]^-$, with 10ppm molecular weight tolerance. The quality of the chromatographic peaks and spectra was inspected, and the plausibility of database candidates was assessed based on retention time, isotope pattern, adduct formation and neutral losses. Identification was confirmed by reanalysis of representative samples from pancreatic or liver cancer studies and pure chemical standards when available, and comparison of the retention times and the MS/MS spectra. When standards were not available, MS/MS spectra were acquired when possible and compared against those in mzCloud (www.mzcloud.org) or METLIN. The level of identification was determined as proposed by Sumner et al[3]. Further details including the spectra and chromatograms are provided in another supplement to this paper.

**Supplement Methods: Details of metabolite identification**

**General notes:**

Chromatograms and spectra are from representative study samples used as the replication dataset I (EPIC) and from the analysis of pure chemical standards (2-Hydroxy-3-methylbutyric acid: MetaSci, Inc., Toronto, Canada; Ethyl glucoside (A127673): AmBeed, Inc. Arlington Hts, IL, USA).

- Chromatograms and isotope patterns: Find Compounds by Formula in Agilent MassHunter Qualitative Analysis B.06.00 SP1
- Isotope patterns: Red rectangles represent an isotope pattern calculated from the elemental composition (shown in the title of each spectra) , bars inside the rectangles show the observed isotope peaks
- MS/MS spectra: Precursor ion is indicated with a blue dot above the ion. Collision energy is on top of the spectra, e.g. CID@20.0

### 2-Hydroxy-3-methylbutyric acid (HMDB0000407):

#### Negative polarity:

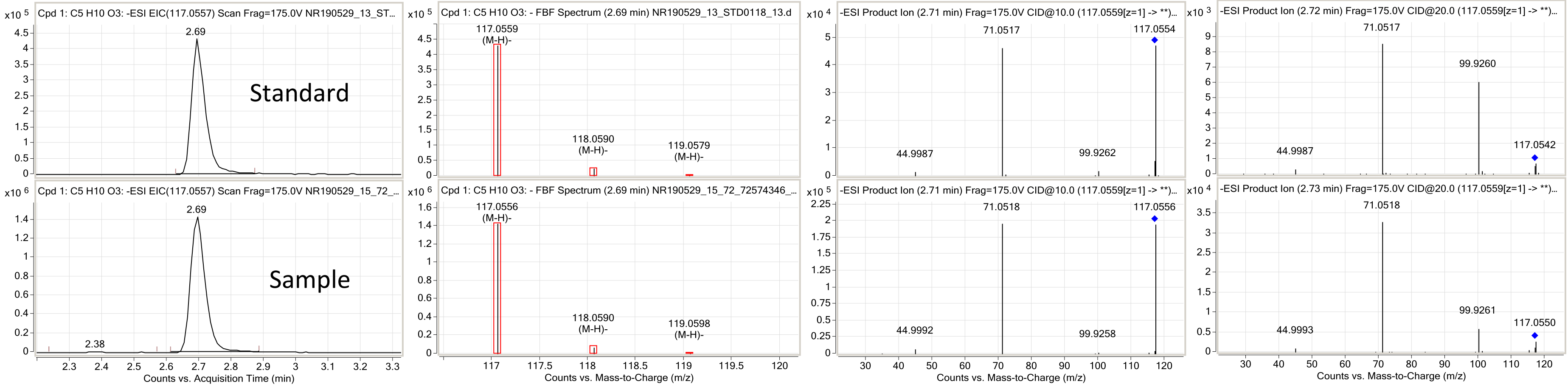

#### Positive polarity:

The compound was found as six related features *m/z* 250.0134, 235.0479, 221.0605, 217.9895, 218.9958, and 203.0227 that are also visible below. MS/MS was performd from the ion of greatest intensity (*m/z* 203.0227). The ion *m/z* 141.053 is $[M+Na]^+$.

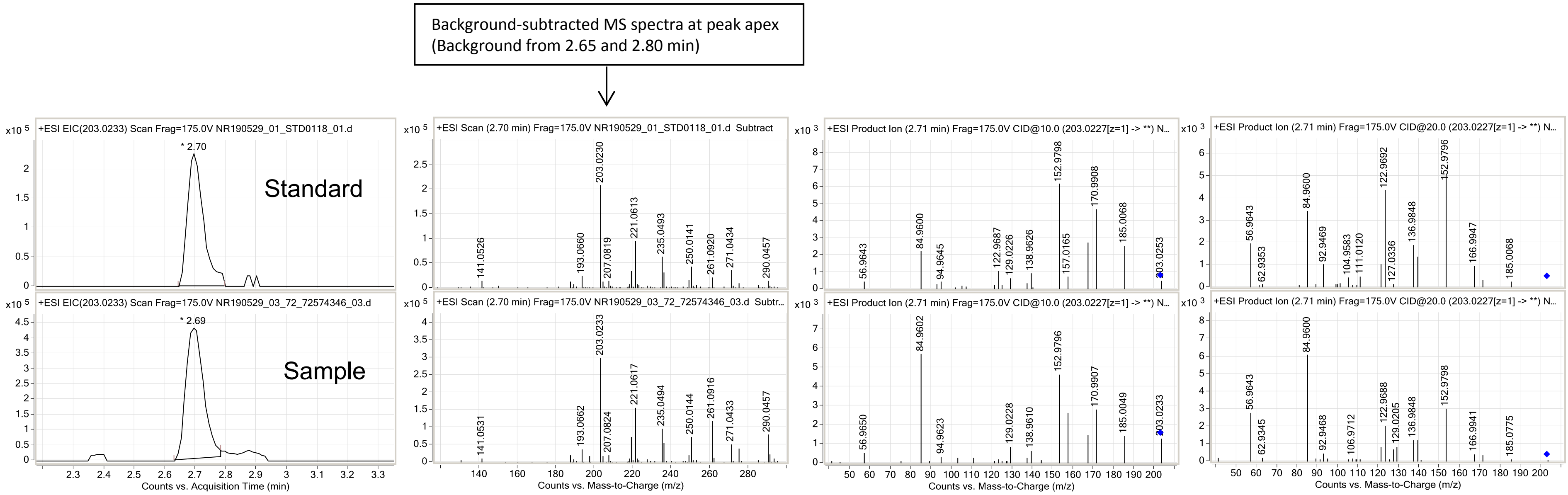

**Unknown** (suspected combination of ethyl-α-D-glucoside, ethyl-β-D-glucoside, and an isomer):

**Positive polarity:**

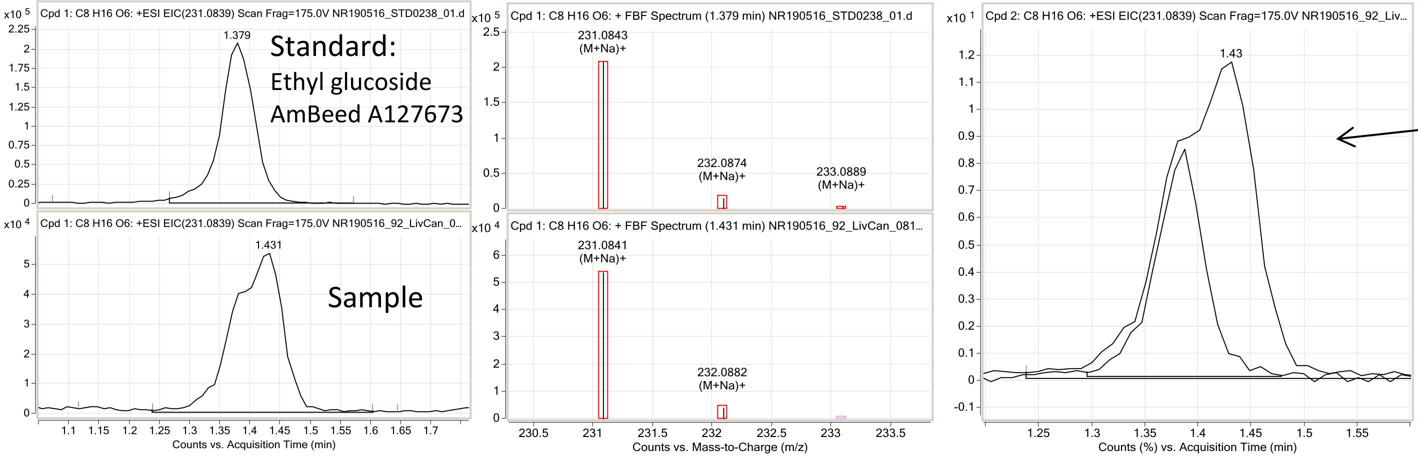

Overlaid chromatograms of the standard and unknown analyzed within the same run.

The unknown appears to be an unresolved peak of two isomers of ethyl glucoside (Mishima *et al*. Biosci Biotechnol Biochem 72 (2008) 393, and an additional unknown isomer of ethyl glucoside.

The standard A127673 was confirmed by AmBeed Inc to be a racemic mixture.

**Negative polarity:**

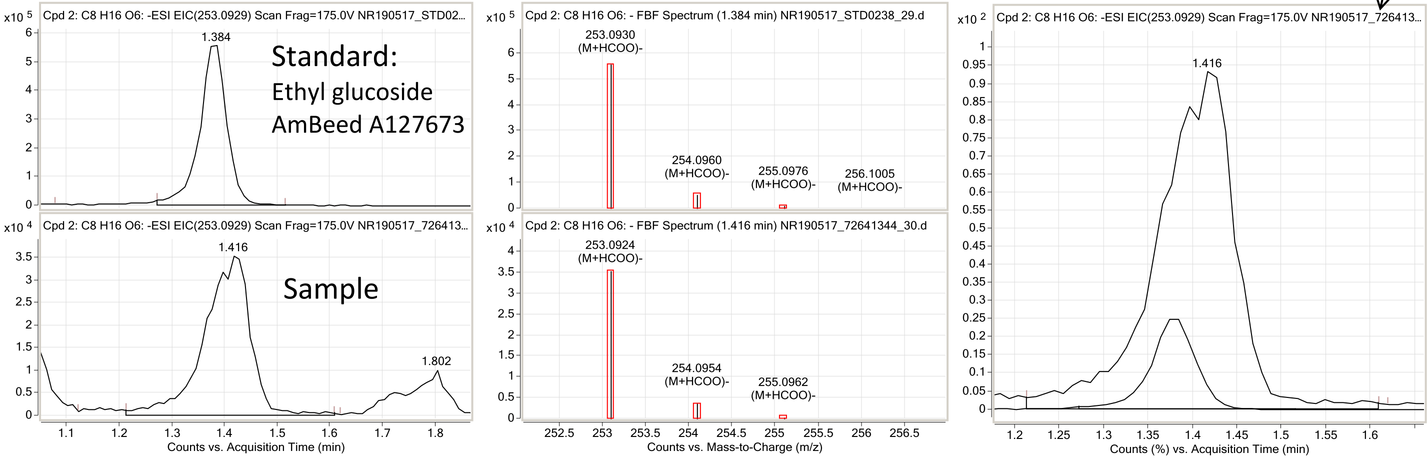
